# ASSESSMENT AND CHARACTERIZATION OF COVID-19 RELATED COGNITIVE DECLINE: RESULTS FROM A NATURAL EXPERIMENT

**DOI:** 10.1101/2023.11.06.23298101

**Authors:** Zennur Sekendiz, Sean A. P. Clouston, Olga Morozova, Melissa A. Carr, Ashley Fontana, Nikhil Mehta, Alina Ali, Eugene Jiang, Benjamin Luft

## Abstract

**Background:** Cognitive impairment is the most common and disabling manifestation of post-acute sequelae of SARS-CoV-2. There is an urgent need for the application of more stringent methods for evaluating cognitive outcomes in research studies.

**Objective:** To determine whether cognitive decline emerges with the onset of COVID-19 and whether it is more pronounced in patients with Post-Acute Sequelae of SARS-CoV-2 or severe COVID-19.

**Methods:** This longitudinal cohort study compared the cognitive performance of 276 patients with COVID-19 to that of 217 controls across four neuroinflammation or vascular disease-sensitive domains of cognition using data collected both before and after the pandemic starting in 2015.

**Results:** The mean age of the COVID-19 group was 56.04±6.6 years, while that of the control group was 58.1±7.3 years. Longitudinal models indicated a significant decline in cognitive throughput ((*β*=-0.168, *P*=.001) following COVID-19, after adjustment for pre-COVID-19 functioning, demographics, and medical factors. The effect sizes were large; the observed changes in throughput were equivalent to 10.6 years of normal aging and a 59.8% increase in the burden of mild cognitive impairment. Cognitive decline worsened with coronavirus disease 2019 severity and was concentrated in participants reporting post-acute sequelae of SARS-CoV-2.

**Conclusion:** COVID-19 was most likely associated with the observed cognitive decline, which was worse among patients with PASC or severe COVID-19. Monitoring patients with post-acute sequelae of SARS-CoV-2 for declines in the domains of processing speed and visual working memory and determining the long-term prognosis of this decline are therefore warranted.

## Introduction

The World Health Organization and the Centers for Disease Control and Prevention use different criteria to define PASC as a condition characterized by persistent Coronavirus Disease 2019 (COVID-19) symptoms: the former requires symptoms to appear three months after the SARS-CoV-2 infection (COVID-19) onset and last for at least two months, while the latter requires at least four weeks of symptom persistence with no other explanation. [1] [1, 2, 3]

Cardiopulmonary such as dyspnea and chest pain, and neuropsychiatric symptoms such as brain fog, fatigue, headache, and depression were reported to be the most frequent complaints of PASC patients.[2] Survivors of COVID-19 were found to seek more frequent medical care for respiratory, diabetes, and neuropsychiatric disorders at 6 months after the onset of SARS-CoV-2 infection compared to their pre-COVID-19 baseline.[3]

Cognitive impairment has been reported as a prevalent and incapacitating condition among patients reporting the presence of post-acute sequelae of SARS-CoV-2 (PASC) [4, 5] that can persist for more than 12 weeks after COVID-19 symptom onset.[6] Reported risk factors for COVID-19-related cognitive decline (CRCD) include common risk factors for age-related dementia including older age, more severe acute COVID-19 severity, female gender, educational attainment, angiotensin-converting enzyme 2 receptor overexpression, social isolation-related psychiatric symptoms, gut dysbiosis and dementia risk factors such as obesity, diabetes, cardiovascular disease, and hypertension. Intriguingly, COVID-19 vaccination can reduce the risk of CRCD both before and after contracting COVID-19,[4, 7] potentially via the removal of latent infections and the restoration of normal function of the immune and inflammatory systems.

Executive dysfunction is commonly reported following many infections including but not limited to West Nile virus, Human Immunodeficiency, Hepatitis C, and Chikungunya viruses.[8] CRCD also appears to manifest as executive dysfunction and can include changes to working memory, verbal fluency, attention, and processing speed.[5, 9] Consistent with a view that COVID-19 might cause CRCD, executive dysfunction is associated with COVID-19 severity, [4] and is evident in both mild and moderate COVID-19,[10] irrespective of age at infection and may continue up to two years after COVID-19.[3, 11] Additionally, acute and subacute infarctions are the most common brain MRI findings in studies of neurological PASC, followed by olfactory bulb abnormalities, white matter abnormalities, cerebral microbleeds, and gray matter abnormalities.[13] However, CRCD might manifest predominantly as subjective impairments, with difficulties in inattention, forgetfulness, and brain fog [4] being frequently reported alongside other commonly presenting neurological symptoms such as headaches, fatigue, dizziness, sleep-related symptoms, and ageusia/anosmia.[12] Yet, while subjective cognitive symptoms are important to functional limitations, they only mildly concur with changes to cognitive performance.[14]

Despite growing concerns that PASC may cause cognitive impairment, existing research lacks control groups or pre-COVID-19 cognitive assessments.[15] As cognitive impairment is a common comorbidity of PASC and may qualify people for accommodations and disability payments under the Americans with Disabilities Act, a recent viewpoint suggests more thorough studies of cognitive outcomes.[16] Additionally, it remains unclear whether cognitive abnormalities result from SARS-CoV-2 infection or instead reflect pre-COVID-19 functioning that enhances susceptibility to severe COVID-19. In this study, we used a natural experiment that occurred when some essential workers were exposed to COVID-19 while others were not. We hypothesized that cognitive decline would correspond to SARS-CoV-2 infection and would be more severe in patients with more severe acute COVID-19 or PASC.

## Methods

### Ethics Statement

Informed written consent was obtained from all participants or their surrogates, and no financial compensation was provided. Herein, we follow the STROBE reporting guidelines for cohort studies. This study was approved by the Stony Brook University Institutional Review Board (CORIHS-A, IRB#604113).

### Participants

Participants who underwent computer-assisted testing on a neuropsychological battery between 11/2015-12/2019 and had at least one follow-up collected between 3/2020-2/2023 for an occupation-based study of cognitive aging were eligible for the present study. Cohort participants who reported having COVID-19 symptoms and a positive COVID-19 polymerase chain reaction (PCR), antibody, or antigen test result between 3/2020 and 11/2021 were included in the COVID-19 infected group. The “verified” COVID-19 group included patients who provided proof of SARS-CoV-2 infection with a positive COVID-19 antigen, antibody, or PCR test. The “unverified” COVID-19 group included participants who reported positive laboratory test findings but for whom we were unable to gather the recorded evidence. Participants with multiple sclerosis, Parkinson’s disease, dementia, or stroke were excluded.

The COVID-19 group was separated into two subgroups based on persistent symptoms. Participants who reported experiencing at least one of the COVID-19-related symptoms lasting ≥4 weeks were placed in the PASC group, while those who did not experience any such symptoms were placed in the non-PASC group. The duration of persistent COVID-19-related symptoms ranged from ≥4 weeks with unspecified duration to more than 3 years from the onset of COVID-19. A total of 20 participants with COVID-19 lacked PASC status information.

The uninfected (control) group reported that, to the best of their knowledge, they had not had COVID-19. This included neither experiencing COVID-19 symptoms nor receiving a positive PCR, antigen, or antibody test result during the specified timeframe.

Completed vaccination (C) was defined as finishing a vaccine series with a brand-specific dosage schedule. The incomplete group (I) had pending vaccinations. In the COVID-19 group, we further stratified vaccination status by the time of vaccination relative to SARS-CoV-2 infection.

### Data Collection

Electronic medical records were used to gather clinical data, and the diagnoses of heart disease, diabetes, hypertension, depression, and hyperlipidemia were done by healthcare providers. BMI was calculated as weight (kg)/ [height (m)]^2^.

Initial COVID-19 symptoms, vaccination information, hospitalization (defined as a hospital stay of ≥24 h), and diagnostic data were gathered through a self-reported survey, text messages, phone interviews, follow-up visits, and data from medical records outside the study.[17] Self-reported demographic data were obtained during registration or the initial visit and were updated during follow-up visits. The analysis used the most recent data collected before March 1, 2020.

### COVID-19 Severity

Acute COVID-19 was classified into three categories: mild, moderate, and severe, according to the October 2021 NIH COVID-19 clinical spectrum guidelines.[17] Asymptomatic cases were excluded from the present study.

### PASC Symptoms

PASC symptoms were classified into the following categories: Respiratory; dyspnea, sore throat, congestion, runny nose, wheezing, and cough; Cardiac; chest pain and palpitations, Neurological symptoms: Central Nervous System (CNS); dizziness, vertigo, brain fog, lethargy, tinnitus, headache; Peripheral Nervous System (PNS); loss of smell and taste, pins and needles; Psychiatric: anxiety, depression, and post-traumatic stress disorder (PTSD) Muscular; body aches and pains, and joint pain; Gastrointestinal: nausea, vomiting, diarrhea, and weight loss; Fatigue; “very tired”; “low energy” Others included fever and rash.

### Cognitive Function Assessment

The ultra-sensitive Cog State Brief Battery, a computerized neuropsychological examination, was used to assess cognition.[18] Cog State detects modest cognitive abnormalities over repeated evaluations and is sensitive to dementia. It assesses cognitive performance using three game-like tasks (detection, identification, and one-card learning) that include repeated trials using a green-background virtual deck of playing cards. Participants were given instructions and then played games using two keyboard keys (marked “Y” for yes and “N” for no).[19] The test was used to measure four metrics that are sensitive to neuroinflammation or vascular disease and influence executive function: response speed, processing speed, cognitive throughput, and visual working memory. Reaction speed measures the detection task completion rate (answers per second). Processing speed (answers per second) measures the average number of correct answers in identification tasks.[20] Throughput measures one-card learning accuracy divided by testing speed (correct answers per second). Visual memory was measured using accuracy on a one-card learning task.

Executive function domains include the ability to inhibit impulsive responses (inhibition), to store information in working memory while other cognitive processes are occurring (working memory), to switch attention quickly from one stimulus or set of rules to another (switching), and to track novel stimuli and replace old information (updating).[21] The lack of a standardized definition for executive function (EF) poses challenges in accurately assessing its functionality.[22] One of the four domains we looked at here that influence executive function is processing speed, which may cause people with slower processing speed to have trouble activating executive or decision-making processes in time before being provided with alternative information or task demands.[21] Viral infections can indirectly impact episodic memory by reducing the attention and processing speed necessary for encoding.[23]

Cognitive exams were done during annual monitoring visits; therefore, the timing of the test administration differed for each participant. Participants included in this analysis had ≥2 assessments during the study period, with ≥1 before and ≥1 after the onset of COVID-19 symptoms. In the uninfected group, we estimated a counterfactual symptom onset date using the mean date of onset for those infected. Details of the timing of cognitive assessments across groups are shown in Figure 2.

### Statistical Analysis

The means of categorical variables were compared using the Chi-square test, and those of continuous variables were compared using the Student’s t-test.

This study represents a secondary data analysis of the onset of an unexpected event during study completion. Post hoc power analyses using simulations accounting for moderate intra-individual correlation in the outcome suggested that this study would have power = 0.93 to detect a longitudinal change in throughput with a moderate (D>0.2) effect size.[24]

Linear longitudinal mixed models were used to examine the rate of cognitive decline and detect evidence of change after an acute SARS-CoV-2 infection. We used the date of COVID-19 symptom onset to detect changes across six measures of cognition by comparing trajectories of functioning pre- and post-COVID-19 using a longitudinal discontinuity design.[25] We controlled for individual differences at baseline using random intercepts and estimated the rate of cognitive decline expected after SARS-CoV-2 infection. We projected the expected rate of decline before and after the infection and compared the post-infection decline with the rate among the uninfected cohort. We adjusted for learning effects using an indicator based on a participant’s initial assessment of this cognitive battery. We also incorporated a covariate identifying the size of the difference between those who were infected with SARS-CoV-2 and those who were not to determine the extent of pre-infection differences. We did not model post-infection accelerations in cognitive decline because the annual monitoring schedule did not provide sufficient time points following infection to permit reliable estimates.

Secondarily, we examined variations in COVID-19 severity and the presence of any PASC as potential moderators of the cognitive trajectories. We computed three effect-size estimates: First, we translated regression coefficients, which differ by the scales of the outcome variables, into standardized regression coefficients (labeled “*b*”). Second, we estimated “age-equivalent years (AEY)” as the number of years of normal cognitive aging necessary to cause similar levels of cognitive decline reflected in changes attributed to COVID-19. AEY was calculated by dividing the coefficient for the COVID-19 indicator by the coefficient of the slope attributed to age in the same model. Third, we used descriptive information alongside model-derived information to estimate the expected proportion of participants who would qualify as having a new-onset mild cognitive impairment (NOMCI).[26] at their initial post-COVID-19 visit. We did not require complete cases to evaluate change over time but instead incorporated data from any individual who fit the eligibility criteria. Missing data in longitudinal models is handled more effectively using the EM algorithm in longitudinal multilevel modeling[27] than by using multivariate imputation if we can assume data are missing at random or for reasons unrelated to the participant’s average rate of decline over time. Models were adjusted for the possible confounding variables, including age, gender, race, BMI, education, depression, diabetes, hypertension, hyperlipidemia, and cardiovascular diseases. We included vaccination status and vaccination timing relative to the timing of infection in the descriptive analysis but not in the regression analysis due to the small number of participants who were fully vaccinated before the onset of infection. All analyses were performed in STATA-17/MP [StataCorp].

## Results

### Participants

After the application of the inclusion and exclusion criteria, we analyzed data from a total of 493 eligible participants (Figure 1). As shown in Table 1, the infected and uninfected groups had similar demographics and health conditions before infection, though those who reported infections were somewhat younger (mean age=58.1 years) as compared to 56.0 years in the COVID-19 group (*P*=.001). Both groups were made up of Caucasian men, and a majority had at least some college education. Only 19 subjects with COVID-19 had immunization at the onset of infection. A subgroup of 46.6% (N=119) of COVID-19 survivors reported PASC. The PASC group had more cardiac disease before COVID infection than those without PASC (Table 2). Chronic fatigue (42.0%), Central Nervous System (CNS) (40.3%), and respiratory symptoms (36.9%) were the most common PASC symptoms. Brain fog included 95% of CNS symptoms some with other co-existing neurological symptoms.

**Figure 1.**
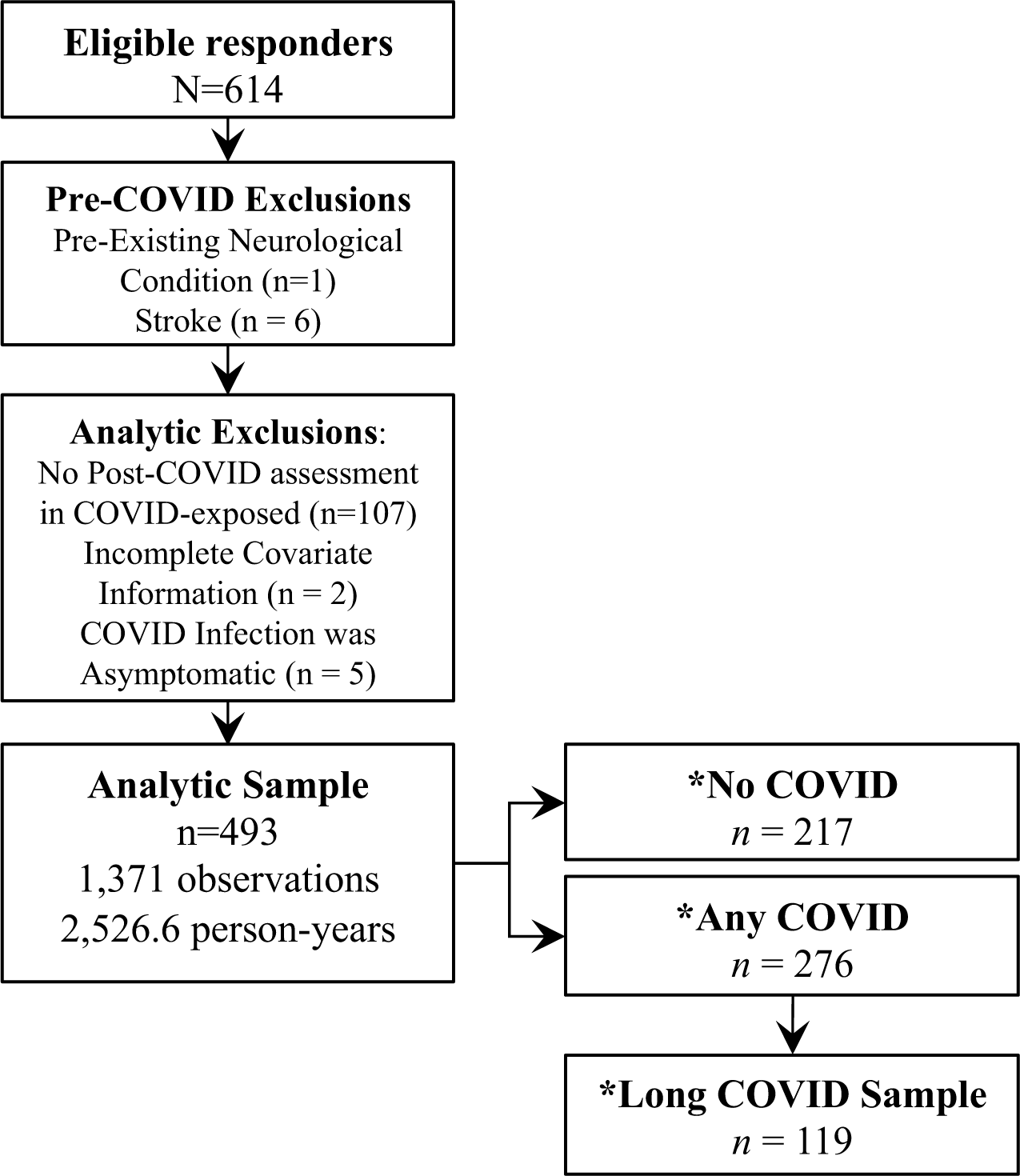
Sample Inclusion and Exclusion Criteria

**Table 1.**
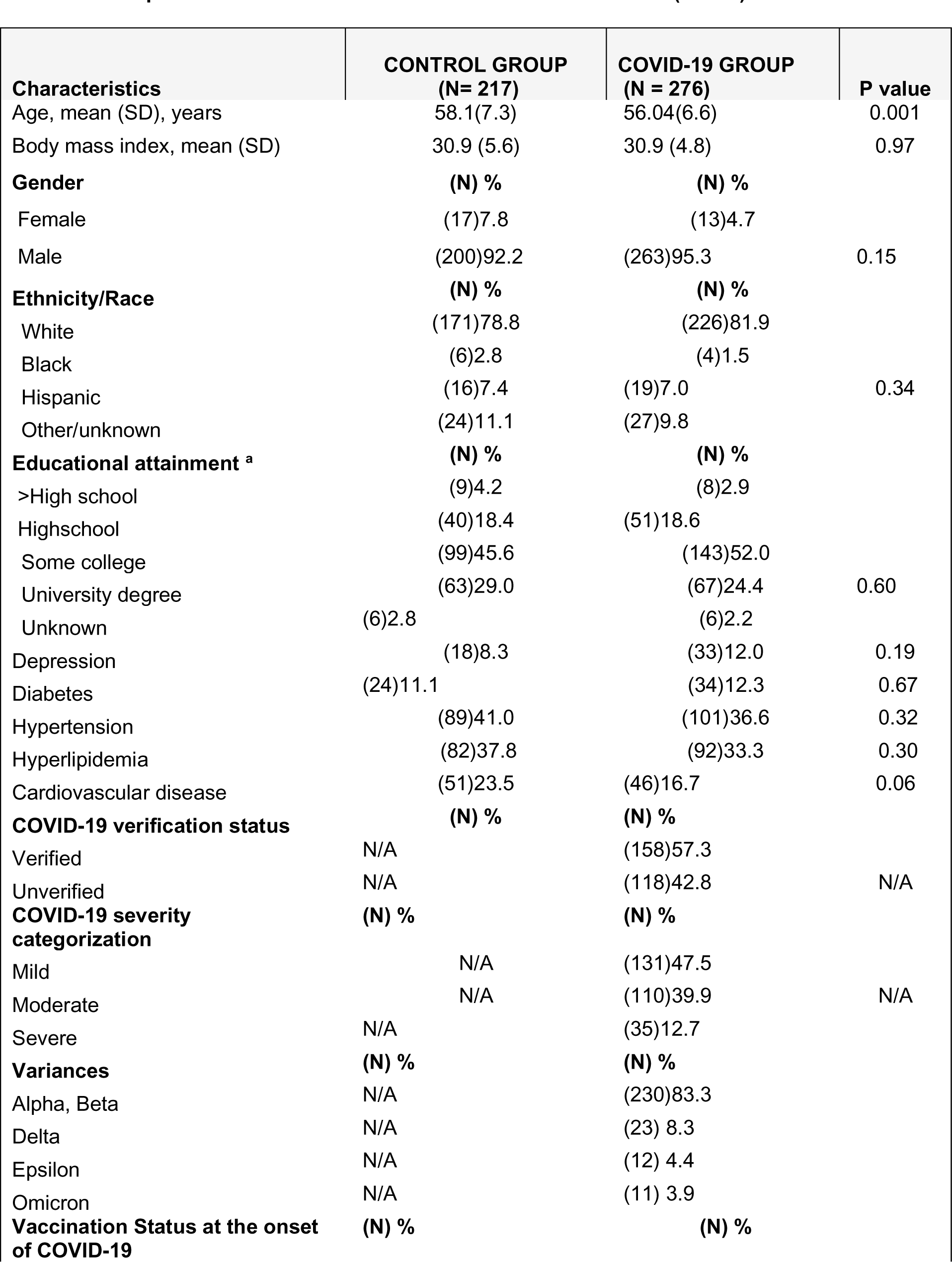
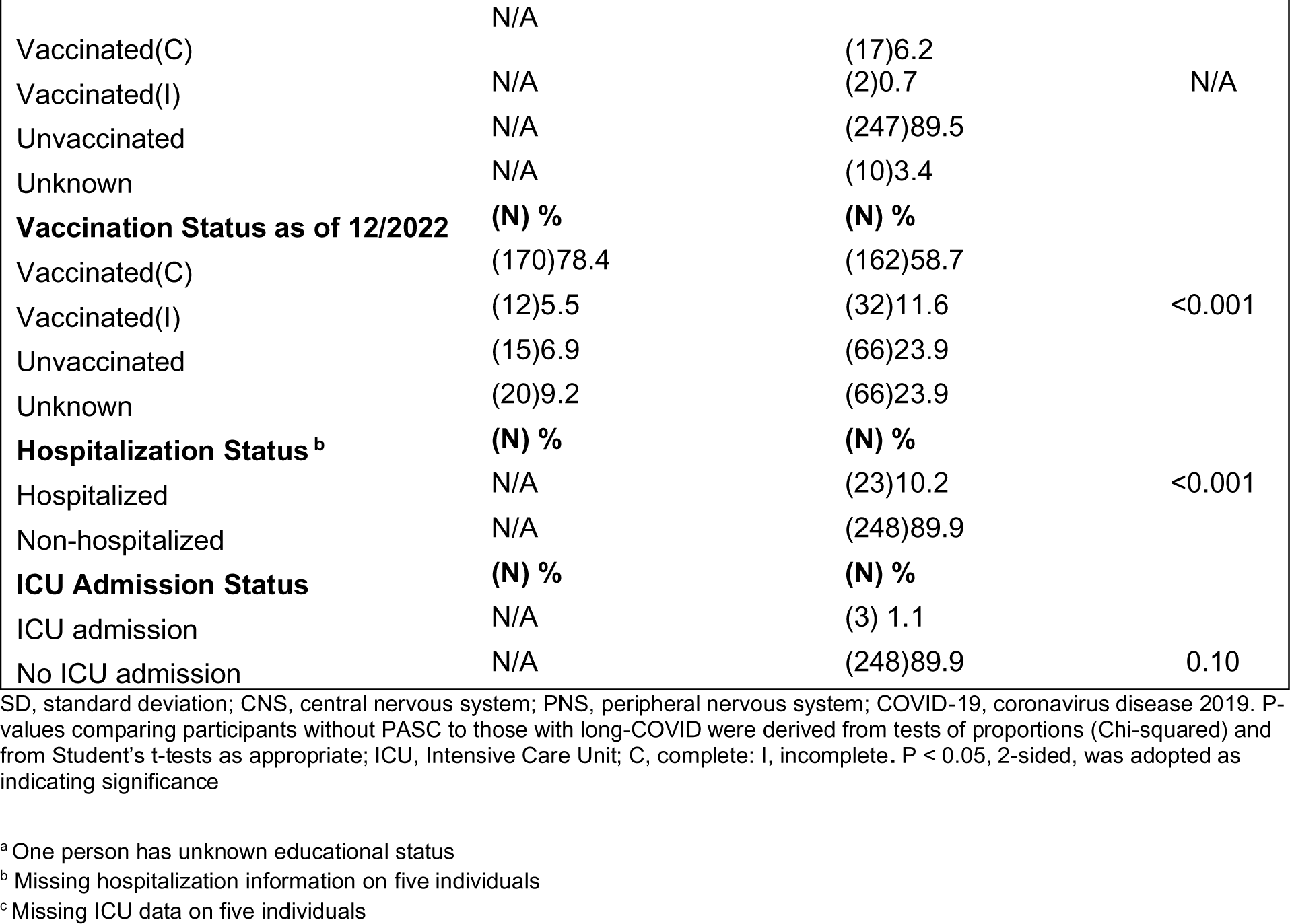
Participant Characteristics: Without Versus With COVID-19. (n=493) ^a^.

**Table 2.** Responder Characteristics: Without Versus With Post-acute Sequelae of SARS-CoV-2 (PASC) (n=256) ^a^.

| **Characteristics** | **Non-PASC (n = 137)** | **PASC (n = 119)** | **P value** |
| --- | --- | --- | --- |
| Age, mean (SD), years | 55.71 (6.48) | 56.34 (7.11) | 0.46 |
| Body mass index, mean (SD) | 30.68 (4.88) | 31.22 (4.55) | 0.36 |
| **Gender** | **(N) %** | **(N) %** |  |
| Female | (7)5.11 | (5)4.20 |  |
| Male | (130) 94.89 | (114)95.80 | 0.73 |
| **Ethnicity/Race <sup>b</sup>** | **(N) %** | **(N) %** |  |
| White | (111) 81.02 | (98)82.35 |  |
| Black | (4)2.92 | (0)0.00 |  |
| Hispanic | (9)6.57 | (9)7.56 | 0.34 |
| Other/unknown | (13)9.49 | (12)10.08 |  |
| **Educational attainment <sup>c</sup>** | **(N) %** | **(N) %** |  |
| >High school | (3)2.19 | (5)4.24 |  |
| Highschool | (26)18.98 | (20)16.95 |  |
| Some college | (68)49.64 | (62)52.54 |  |
| University degree | (36)26.28 | (29)24.58 | 0.50 |
| Unknown | (4)2.92 | (2)1.69 |  |
| Depression(N) % | (12)8.76 | (19)15.97 | 0.08 |
| Diabetes(N) % | (13)9.49 | (17)14.29 | 0.23 |
| Hypertension(N) % | (43)31.39 | (50)42.02 | 0.08 |
| Hyperlipidemia(N) % | (39)28.47 | (46)38.66 | 0.08 |
| Cardiovascular disease(N) % | (17)12.41 | (27)22.69 | 0.03 |
| **Lingering symptoms type <sup>d</sup>** | **(N) %** | **(N) %** |  |
| CNS | N/A | (48)40.34 | N/A |
| PNS | N/A | (19)15.97 | N/A |
| Muscular | N/A | (27)22.69 | N/A |
| Psychiatric | N/A | (8)6.72 | N/A |
| Fatigue | N/A | (50)42.02 | N/A |
| Gastrointestinal | N/A | (6) 5.04 | N/A |
| Respiratory | N/A | (44)36.97 | N/A |
| Other | N/A | (4)3.36 | N/A |
| **COVID-19 verification status** | **(N) %** | **(N) %** |  |
| Verified | (79) 57.66 | (78)65.55 |  |
| Unverified | (58)42.34 | (41)34.45 | 0.20 |
| **COVID-19 severity categorization** |  |  |  |
| Mild | (75) 54.74 | (42)35.29 |  |
| Moderate | (57) 41.61 | (48)40.34 | <0.001 |
| Severe | (5) 3.65 | (29)24.37 |  |
| **Vaccination Status at the onset of COVID-19** | **(N) %** | **(N) %** |  |
| Vaccinated(C) | (5)5.15 | (7) 5.88 |  |
| Vaccinated(I) | (1)0.0 | (1)0.84 | 0.35 |
| Unvaccinated | (125)90.44 | (108)90.75 |  |
| Unknown | (6)4.41 | (3)2.52 |  |
| **Vaccination Status as of 12/2022** | **(N) %** | **(N) %** |  |
| Vaccinated(C) | (85) 63.04 | (69) 57.98 |  |
| Vaccinated(I) | (12) 8.76 | (12) 10.98 | 0.69 |
| Unvaccinated | (30) 21.90 | (32)26.89 |  |
| Unknown | (10)7.30 | (6)5.04 |  |
| **Hospitalization Status <sup>e</sup>** | **(N) %** | **(N) %** |  |
| Hospitalized | (2)1.46 | (21)21.85 | <0.001 |
| Non-hospitalized | (132) 96.35 | (97)78.15 |  |
| **ICU Admission Status** | **(N) %** | **(N) %** |  |
| ICU admission | (0) 0.0 | (3)1.92 |  |
| No ICU admission | (137)100 | (116) 98.08 | 0.10 |

SD, standard deviation; CNS, central nervous system; PNS, peripheral nervous system; COVID-19, coronavirus disease 2019. P-values comparing participants without PASC to those with long-COVID were derived from tests of proportions (Chi-squared) and from Student's t-tests as appropriate; ICU, Intensive Care Unit; C, complete; I, incomplete. P < 0.05, 2-sided, was adopted as indicating significance
<sup>a</sup>Among 276 COVID-19 group, 20 participants had unknown PASC status
<sup>b</sup>Other; included participant who declined to specify race or ethnicity
<sup>c</sup>One person has unknown educational status
<sup>d</sup>Some participants reported more than 1 symptoms
<sup>e</sup>missing hospitalization information on PASC (1) and Non-PASC (3) groups

**Table 3.** Participants self-reported duration of symptoms after the onset of COVID-19.

| Symptoms duration | Frequency (N=119) | Percentage (%) |
| --- | --- | --- |
| 4 weeks <sup>a</sup> | 10 | 8.40 |
| 1-2 months | 18 | 15.13 |
| 2-6 months | 32 | 26.89 |
| 6-12 months | 18 | 15.13 |
| Greater than 1 year | 16 | 13.45 |
| Greater than 2 years | 17 | 14.29 |
| Greater than 3 years | 8 | 6.72 |

Abbreviations: COVID -19, corona virus disease 2019
<sup>a</sup> Patient self-reported in the survey at least 4 weeks of persistent symptoms but did not specify the duration

**Table 4.** Comparison of Responders Who Were and Were not Included in Analysis Based on the availability of PASC status.

| **Characteristics** | **Not included (20)** | **Included (n = 256)** | **P value** |
| --- | --- | --- | --- |
| Age, mean (SD), years | 56.55 (4.59) | 56.00 (6.77) | 0.624 |
| Body mass index, mean (SD) | 30.06 (5.42) | 30.94 (4.73) | 0.430 |
| **Gender** | **(N) %** | **(N) %** |  |
| Female | (1)5.0 | (12)4.69 |  |
| Male | (19) 95.00 | (244)95.31 | 0.95 |
| **Ethnicity/Race <sup>a</sup>** | **(N) %** | **(N) %** |  |
| White | (17) 85.00 | (209)92.48 |  |
| Black | (0)0.0 | (4)1.56 |  |
| Hispanic | (1)5.00 | (18)7.03 | 0.929 |
| Other/unknown | (2)10.00 | (25)9.77 |  |
| **Educational attainment <sup>b</sup>** | **(N) %** | **(N) %** |  |
| >High school | (0)0.0 | (8)3.14 |  |
| Highschool | (5)25.00 | (46)18.04 |  |
| Some college | (13)65.00 | (130)50.98 |  |
| University degree | (2)10.00 | (65)25.49 | 0.391 |
| Unknown | (0)0.00 | (6)2.35 |  |
| Depression(N) % | (2)10.00 | (31)12.11 | 0.779 |
| Diabetes(N) % | (4)20.00 | (30)11.72 | 0.277 |
| Hypertension(N) % | (8)40.00 | (93)36.33 | 0.743 |
| Hyperlipidemia(N) % | (7)35.00 | (85)33.20 | 0.869 |
| Cardiovascular disease(N) % | (2)10.00 | (44)17.19 | 0.406 |
| **COVID-19 verification status** | **(N) %** | **(N) %** |  |
| Verified | (1) 5.0 | (157)61.33 |  |
| Unverified | (19)95.00 | (99)38.67 | <0.001 |
| **COVID-19 severity categorization** | **(N) %** | **(N) %** |  |
| Mild | (14) 70.00 | (117)45.70 |  |
| Moderate | (5) 25.00 | (105)41.02 | <0.105 |
| Severe | (1) 5.00 | (34)13.28 |  |
| **Vaccination Status at the onset of COVID-19** | **(N) %** | **(N) %** |  |
| Vaccinated(C) | (2)10.00 | (11) 4.31 |  |
| Vaccinated(I) | (3)15.00 | (3)1.18 | <0.001 |
| Unvaccinated | (14)70.00 | (233)91.37 |  |
| Unknown | (1)5.00 | (8)3.14 |  |
| **Vaccination Status as of 12/2022** | **(N) %** | **(N) %** |  |
| Vaccinated(C) | (8) 40.00 | (154) 60.16 |  |
| Vaccinated(I) | (8) 40.00 | (24) 9.38 | <0.001 |
| Unvaccinated | (4) 20.00 | (62) 24.22 |  |
| Unknown | (0) 0.00 | (16) 6.25 |  |
| **Hospitalization Status <sup>c</sup>** | **(N) %** | **(N) %** |  |
| Hospitalized | (2) 5.0 | (21) 8.98 | <0.001 |
| Non-hospitalized | (19) 95.0 | (229) 89.45 |  |
| **ICU Admission Status <sup>d</sup>** | **(N) %** | **(N) %** |  |
| ICU admission | (0) 0.0 | (3) 1.17 |  |
| No ICU admission | (0) 0.0 | (249) 98.83 | N/A |

SD, standard deviation; CNS, central nervous system; PNS, peripheral nervous system; COVID-19, coronavirus disease 2019. P-values comparing participants without PASC to those with long-COVID were derived from tests of proportions (Chi-squared) and from Student's t-tests as appropriate; ICU, Intensive Care Unit; C, complete; I, incomplete. P < 0.05, 2-sided, was adopted as indicating significance
<sup>a</sup> Other; included participant who declined to specify race or ethnicity
<sup>b</sup> One person has unknown educational status
<sup>c</sup> Missing hospitalization information on four individuals
<sup>d</sup> Missing ICU data on four individuals

**Table 5.** Coronavirus disease 2019 (COVID-19) Severity Categorization.

| Category | Description |
| --- | --- |
| Asymptomatic | Individuals who test positive for SARS-CoV-2 using a virologic test (i.e., a nucleic acid amplification test [NAAT] or an antigen test) but who have no symptoms that are consistent with COVID-19. |
| Mild | At least one of the following symptoms: fever, fatigue, headache, cough, chills, muscle aches, chest pain, sore throat, loss of smell/taste, N/V/D, congestion, or runny nose<br>Do not have shortness of breath or difficulty breathing.<br>Managed in an ambulatory setting or at home through telemedicine or telephone visits. |
| Moderate | Shortness of breath and/or evidence of lower respiratory disease during clinical assessment or imaging<br>or<br>found to have SpO<sub>2</sub> ≥94% in room air at sea level. |
| Severe | SpO<sub>2</sub> < 93% in room air<br>and/or<br>a respiratory rate > 30 breaths/min<br>and/or<br>HR > 100 BPM or self-endorsed tachycardia<br>and/or acute respiratory distress syndrome; septic shock; cardiac dysfunction; an exaggerated inflammatory response in addition to pulmonary disease; severe illness causing cardiac, hepatic, renal, central nervous system, or thrombotic disease;<br>and/or hospital admission, supplemental oxygen use, ICU admission, or death from COVID-19 |

COVID-19, coronavirus disease 2019; SARS-CoV-2, severe acute respiratory syndrome coronavirus 2; ICU, intensive care unit; HR, heart rate; BPM, beats per minute; N/V/D, Nausea/Vomiting/Diarrhea

### Cognitive Function after SARS-CoV-2 infection

Modeling identified slow cognitive aging over time (standardized regression coefficient *β*=-0.253, *P*<.001). However, while we found no evidence of any difference in cognition between the COVID-19 cohort and uninfected controls before or at the point of symptom onset among the infected group (*β*=-0.016, *P*=.751), we found a significant decrement in cognition among those who developed COVID-19 after their infection (Table 6). Changes in throughput (Figure 2) were substantial (*β*=-0.168, *P*=.001), equivalent to an AEY of 10.6 years of normal aging and 18 new cases of MCI (NOMCI=59.8%) in this sample. The results were also statistically significant for visual memory (*β*=-0.150, *P*=.004; AEY=16.51) (Table 6).

**Figure 2.**
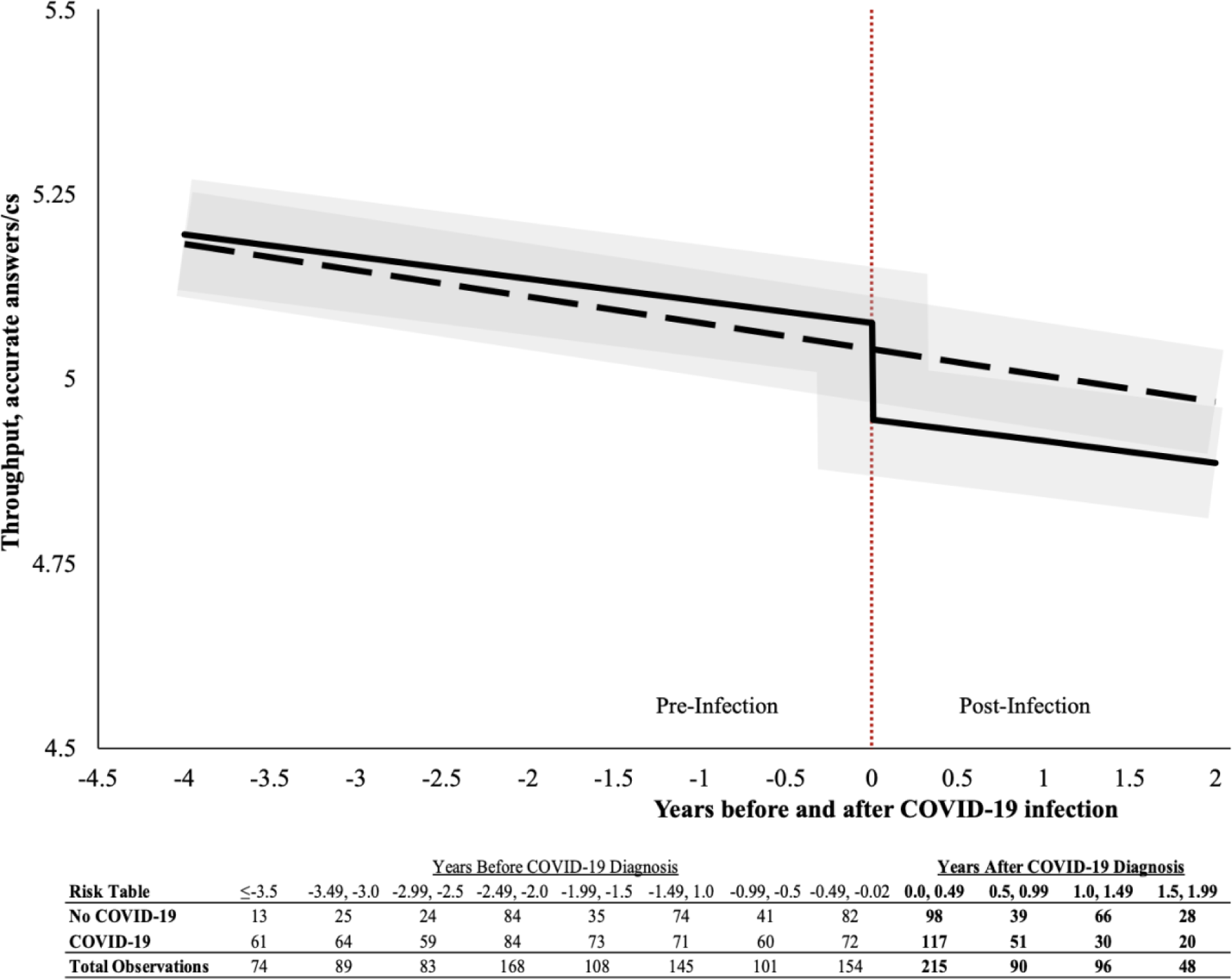
Trajectory plot showing expected throughput before and after the onset of COVID-19 symptoms (time 0). Expectations are stratified as SARS-CoV-2 infected (solid black line) and uninfected (black dashed line). 95% Confidence intervals are shown in translucent gray. The estimated model is *Y*_*it*_ = 5.85 − 0.014 ∗ *A*_*it*_ − 0.146 ∗ *C*_19*t*_[−0.045 ∗ *NC*_19_ − 0.004 ∗ *NC*_19_ ∗ *t*] + ***XB*** + γ_0*i*_ + γ_1*i*_*t* + ε_*it*_, where ***XB*** contains an array of covariates, Ait denotes age of subject i at time t, C19t is a time-varying indicator for the onset of COVID-19 symptoms, and NC19 is a time-invariant indicator of belonging to a COVID-19 cohort: a measurement of selection bias that indicates the differences between individuals who ever developed symptomatic COVID-19 compared to those who were in the comparison group. For the unexposed group, the counterfactual symptom onset time (t=0 in the graph above) was calculated as the average date that those in the COVID-19 group were infected. Results enclosed in square brackets in the equation were not statistically significantly different from 0 but are reflected in the figure anyways. Results for all domains are shown in Table 6. Abbreviations: cs, centi-seconds.

**Table 6.** Longitudinal degree of association between COVID-19 onset versus cognitive performance.

| **Cognitive Domain** | **Std. Coef.** | **Age Eq. Yrs.** | **Coef.** | **Std. Err.** | **P** |
| --- | --- | --- | --- | --- | --- |
| Throughput | **-0.168** | **10.59** | **-0.146** | **0.000** | **0.001** |
| Visual Memory | **-0.150** | **16.51** | **-0.023** | **0.008** | **0.004** |
| Reaction Speed | -0.072 | 6.26 | -0.084 | 0.001 | 0.161 |
| Processing Speed | -0.074 | 6.73 | -0.001 | 0.000 | 0.154 |

**Note:** Models adjusted for age, sex, race/ethnicity, educational attainment, hypertension, body mass, diabetes, post-traumatic stress symptoms, depressive symptoms. $P < 0.05$ , 2-sided, was adopted as indicating significance
Abbreviations: COVID-19: Novel Coronavirus 2019; Std. Coef.: standardized regression coefficient; Age Eq. Yrs.: Number of years of aging necessary to equal a similar level of cognitive decline as evident in COVID-19; Coef.: regression coefficient; Std. Err.: standard error; P: p-value.

### PASC versus Acute COVID-19

Next, we examined whether individuals reporting lingering cognitive symptoms had evidence of different or more severe cognitive decline than those who only reported acute symptoms. Longitudinal results comparing functioning after the onset of COVID-19 to pre-COVID-19 levels (Table 7) suggested cognitive deficits concentrated in PASC cases. While only reaction speed was associated with the onset of acute COVID-19, in participants with PASC, we found significant decrements across all four domains of cognitive performance after the onset of COVID-19, with the largest decrements involving cognitive throughput and response speed.

**Table 7.** Longitudinal associations between COVID-19 onset versus cognitive performance, stratified by presence or absence of Post-Acute Sequelae of COVID-19.

| Cognitive Domain | Acute COVID-19 Only |  |  |  | Post-Acute Sequelae of COVID-19 |  |  |  |
| --- | --- | --- | --- | --- | --- | --- | --- | --- |
|  | Std. Coef. | Coef. | Std. Err. | P | Std. Coef. | Coef. | Std. Err. | P |
| Throughput | -0.070 | -0.687 | 0.506 | 0.175 | **-0.203** | **-2.504** | **0.638** | **<0.001** |
| Visual Memory | -0.043 | -6.771 | 8.066 | 0.401 | **-0.142** | **-32.388** | **11.777** | **0.006** |
| Reaction Speed | **-0.133** | **-1.709** | **0.664** | **0.010** | **-0.188** | **-3.043** | **0.836** | **<0.001** |
| Processing Speed | -0.096 | -0.845 | 0.454 | 0.063 | **-0.121** | **-1.262** | **0.537** | **0.019** |

**Note:** Models adjusted for age, sex, race/ethnicity, educational attainment, hypertension, body mass, diabetes, post-traumatic stress symptoms, depressive symptoms. P < 0.05, 2-sided, was adopted as indicating significance
Abbreviations: COVID-19: Novel Coronavirus 2019; Coef.: regression coefficient; Std.: standardized; Std. Err.: standard error; P: p-value.

### Differences by COVID-19 Severity

Analyses comparing mild with moderate/severe COVID-19 found reduced cognitive throughput and reaction speed compared with pre-COVID-19 functioning in milder COVID-19 cases.

However, more severe COVID-19 cases showed large decrements in cognition across all domains but concentrated in cognitive throughput and processing speed (Table 8).

**Table 8.**
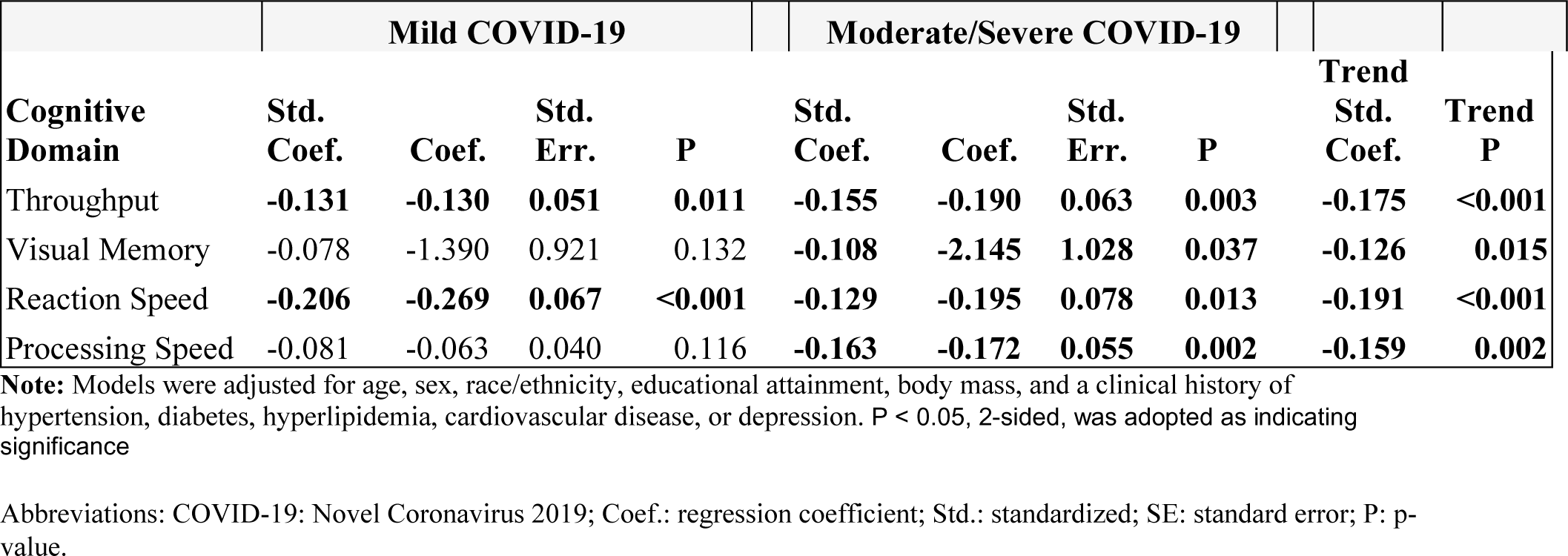
Longitudinal associations between COVID-19 onset versus cognitive performance, stratified by mild and moderate/severe COVID-19.

## Discussion

We aimed to determine whether the development of COVID-19 in a sample of aging essential workers was contemporaneous with decrements in cognitive functioning and whether the declines were steeper in participants with more severe disease and among those reporting PASC. The availability of longitudinal data dating back to 2015 allowed us to investigate whether cognitive decline emerges with SARS-CoV-2 infection or whether the observations reflected lower pre-COVID-19 functioning that rendered patients more susceptible to severe disease, the development of cognitive impairment following the SARS-CoV-2 infection confirmed its association with COVID-19. Among people who experienced symptomatic COVID-19, we found evidence of lasting changes in cognitive functioning equivalent to 10.6 years of normal aging and a 59.8% increase in the number of participants with mild cognitive impairment. These results imply an increase in cognitive dysfunction and poorer brain health after COVID-19, notably in those with more severe disease, or PASC. Patients with severe COVID-19 have been found to have transcriptomic changes in their prefrontal cortex like those of older individuals,[28] and severe COVID-19 yielded a drop in global cognitive performance consistent with decline usually spread over 10 years of normal aging,[29] consistent with the results of our study. According to a large cohort study in a primary care setting, when compared to patients with the diagnosis of acute upper respiratory tract infections (AURI), individuals with the diagnosis of COVID-19 had a significantly higher incidence of mild cognitive disorder (7.6 COVID-19 cases/1,000 person-years vs. 5.1 AURI cases/1,000 person-years), but a lower overall incidence[30] indicating that COVID-19 diagnosed outside of general practice may be misclassified as AURI. This limitation may lower the incidence of COVID-19-related MCI.

Throughput was reduced significantly (*β*=-0.168, *P*=.001) after SARS-CoV-2 infection over at least the following two years for some individuals. We observed no additional decline in throughput in the control group at any point in the same period. This supports the hypothesis that COVID-19 was associated with the cognitive decline observed among people who survived the infection. These results are comparable to those in hospitalized patients with PASC, but they also agree with findings of cognitive impairment in mild COVID-19 cases.[15] We conclude that SARS-CoV-2 infection is most likely associated with cognitive impairment and present the first evidence that cognitive decline emerges with SARS-CoV-2 infection. A Mendelian randomization analysis found that host genetic predisposition to SARS-CoV-2 infection was connected to decreased cognitive function,[31] confirming that COVID-19 infection is a probable trigger of cognitive decline.

Infection severity and cognitive function are well studied, although results are inconclusive.[32] We show here that cognitive performance decreased significantly across measures of executive function, with focal results in subscales of throughput, reaction speed, processing speed, and visual memory that corresponded in time to COVID-19 and were more pronounced in those with greater disease severity. Thus, our findings suggest that differences in cognition among individuals in midlife likely emerged coincident with COVID-19 and likely do not reflect pre-infection differences in cognition.

While only throughput was impacted in individuals without PASC, processing speed, visual memory, and reaction time were similarly impacted in those with PASC. Cognitive deterioration in the non-PASC and PASC groups has been recorded, although the importance of PASC has not been previously studied by specific comparisons.[33] We, therefore, contribute to the existing literature by determining the cognition domains affected and the differential severity of the impairment in PASC versus non-PASC groups.

Slower responses, lower processing speeds, and poorer cognitive throughput and visual memory are domain-specific results of this study, and daily activities require these functions. Response speed is crucial for preventing accidents and falls in older adults, and reduced processing speed and working memory dysfunction are commonly associated with changes in numeracy and poorer financial decision-making in older adults.[34] These domains are thought to represent changes in different cerebral functional systems, with changes in memory often associated with hippocampal dysfunction.[35, 36] In contrast, response and processing speed are usually influenced by white-matter dysfunction although in neuroinflammatory disorders, processing dysfunction may additionally indicate difficulties in coordinating activities in the brain stem and cerebellum.[37] Executive function abnormalities can be caused by lesions in the frontal lobes and other areas of the brain; research suggests that diffuse white matter hyperintensity volume may be more strongly associated with executive function impairments than localized lesions in particular sites.[22] CRCD may thus place affected individuals at risk of health and financial issues; monetary and functional deterioration are common as many as 6 months after hospitalization for COVID-19.[38]

Neurological manifestations of PASC have been attributed to various mechanisms, including endothelial disruption, systemic inflammation, neuroglial dysfunction, SARS-CoV-2 neurotropism, and viral-induced coagulopathy.[39] Several viruses were previously implicated in the ongoing central nervous system damage in humans. Survivors of Herpes Simplex Virus (HSV) Encephalitis had long-term neuropsychiatric symptoms, with their development attributed to cytolysis and inflammation. Alzheimer’s disease (AD) has been linked to a chronic pathogenic mechanism that involves recurrent HSV infections. The human immunodeficiency virus (HIV) is linked to HIV-associated neurocognitive disorders due to persistent Central Nervous System infection (CNS).[40]

Unlike dyspnea and depression, post-viral fatigue and cognitive impairment have no effective therapies. Finding the neurological underpinnings of PASC-related fatigue and cognitive impairment, along with related factors and causes, is crucial due to its possible negative impact on the economy, and its potential to reduce quality of life.[6, 41, 42] Patients, notably those with PASC, should therefore undergo routine screening with tools capable of detecting slight cognitive abnormalities.

Dementia and cognitive decline are only some of the many long-term neurological conditions that have been linked to neurotropic respiratory viruses. COVID-19 may induce serious behavioral issues and call for close neurological and psychiatric monitoring due to the presence of neuroinflammation in many neurodegenerative disorders.[42] In light of the limited availability of evidence regarding COVID-19-related cognitive impairment, the World Health Organization (WHO) has recommended the revision of rehabilitation approaches for related conditions. Additional research is required to effectively implement evidence-based treatment modalities for these individuals.[23]

### Limitations and Strength

We assessed cognitive performance using the highly sensitive Cog State brief battery test and thoroughly measured the cognitive domains that are susceptible to neuroinflammation and vascular alterations. Such self-administered tests advantageously decrease administrator and observer bias. We adjusted our models for most recognized risk variables for cognitive impairment to lessen the confounding effect. By employing a control group with the COVID-19 cases that contained cognitive data from pre/post-COVID-19 on the same patients, we were also able to minimize sampling and information bias. Recent self-reported data on the duration of PASC symptoms collected following cognitive testing helps reduce sampling bias.

Generalizability may be limited because the study population was mostly Caucasian men with a college degree. However, prior studies revealed no correlation between the COVID-19 research findings and conditions related to prior occupational exposures.[17] Thus, the general population with similar characteristics that is not exposed to WTC disaster might benefit from our findings. Even though the majority of our patients contracted COVID-19 when alpha or beta variants were active (83.3%) (Table 1) and therefore contracted the virus before the COVID-19 vaccination campaigns, other studies have suggested that even patients with the Omicron variant reported possible CRCD.[43] Our control group reported no influenza-like symptoms or positive COVID-19 test results during the study period, but they may have been asymptomatic or had false negative results, which would bias the estimated effect sizes towards the null. A small number of participants vaccinated before the SARS-CoV-2 infection prevented us from studying the potential protective effects of vaccination on COVID-19-related cognitive decline.

## Conclusion

Infection with SARS-CoV-2 is associated with cognitive impairment equivalent to 10.6 years of normal aging and a 59.8% increase in MCI. PASC and severe disease appeared to accelerate the cognitive deterioration, especially in executive function. Cognitive impairment affects daily, occupational, and social functioning. Hence, COVID-19 patients, particularly those with PASC, should be evaluated for processing speed and visuospatial ability and treated to prevent cognitive deterioration. Determining the long-term prognosis of the impairments we observed here will be crucial.

## Author Contributions

Concept and design: Sekendiz, Clouston, Luft

Acquisition, analysis, or interpretation of data: All authors.

Original draft, Sekendiz

Critical revision of the manuscript for important intellectual content: Sekendiz, Clouston, Luft, Morozova.

Statistical Analysis: Clouston, Sekendiz

Administrative, technical, and material support: Fontana, Carr

## Data Availability

Data include sensitive private health information. As such, data can be accessed upon receipt of a written request to the corresponding author and the completion of a data use agreement.

## Acknowledgements

We thank World Trade Center Health and Wellness Program patients, staff and faculty for their research contributions.

## Funding

This study was funded by CDC/NIOSH-5 U01OH012275-02-00 (Drs. Luft and Morozova) and CDC-75D30122c15522 (Drs. Luft) and by NIH/NIA R01 AG049953 (Dr. Clouston)

The funders had no part in the study’s design, data collection, analysis, or interpretation, manuscript writing, or publication decision.

Data Sharing Statement, only processed de-identified data will be provided upon written request to the appropriate author due to medical confidentiality.

